# Addiction of Smartphones and Its Relationship to Academic Achievement of Medical Students in Saudi Arabia,2022

**DOI:** 10.1101/2022.12.12.22283066

**Authors:** Rayan Saud Alharbi, Baderldeen Abdulrahman Mohamed, Thamir M Alshammari

**Affiliations:** Ministry of Health, Al Nabhaniyah Hospital, Qassim, Saudi Arabia; College of Applied Medical Sciences, King Saud University, Riyadh, Saudi Arabia; Medication Safety Research Chair, King Saud University, Riyadh, Saudi Arabia

**Author notes:** **Corresponding Author Thamir M. Alshammari, Ph.D, M.S, RPh**, Associate professor of pharmacy practice, Senior Researcher, Medication Safety Research Chair, King Saud University, Riyadh, Saudi Arabia, College of Applied Medical Sciences, King Saud University, Riyadh, Saudi Arabia, Pharmacoepidemiology and Pharmacovigilance Consultant, P.O. Box 2457, Riyadh 11451.

**Keywords:** Smartphones, Awareness, Academic performance, Behavioral intention, Medical Students

## Abstract

**Background:** Smartphones and their increasing capabilities have helped humans to communicate and perform many tasks and it leads to a form of dependency, and it may have negative effects on everyone, especially students.

**Objectives:** To assess smartphone addiction and its relationship to academic performance among medical students at King Saud University, Kingdom of Saudi Arabia.

**Methods:** An observational cross-sectional study was conducted from July to September 2022 including students of the College of Medicine at King Saud University, Kingdom of Saudi Arabia. The data collection tool was structured and utilized an electronic survey.

**Results:** A total of 330 participants answered the study questionnaire. The most common age range of study participants was 18-28 years with 64.2% of the study sample. Male participants represented 63%. The study income is less than 5000 riyals 54.5% per month. Majority of ftudents (65%) believe that using smartphones them to study more efficiently. Analysis of the study results shows that there is a statistically significant correlation between phone addiction and a decrease in the academic performance of college students.

**Conclusion:** Our study found that there is a significant correlation between phone addiction and a drop in academic performance. Despite its attractiveness, smartphone addiction is a time waster for students that might disrupts their sleep and causes stress. It is, therefore, necessary to create a comprehensive plan that directs the students towards balanced use.

## Introduction

Today, the world is transforming in the information and communication field. Smartphones have become a must-have device due to their ability to access information faster. Any problem can solve with one touch, and it is not easy for people to live without it, and it can cause behavior change. The current communication condition is as follows: smartphones are the most well-known. It was created to address everyday accessibility difficulties and has hundreds of built-in apps, and laptops are now required to have an internet connection regularly. Smartphones are advantageous for various reasons, including connectivity and information availability [1]. Smartphones have progressively transformed into indispensable educational tools for anyone. They have evolved into an essential educational tool for all students since they allow them to access school materials via e-learning (Blackboard) and obtain any information [2,3], as well as surf the Internet, entertain themselves, and interact. It has become a crucial part of our daily lives, and this trend is expected to continue. Experts, on the other hand, are increasingly emphasizing its drawbacks, such as the disintegration of social bonds [4]. Medical students and junior physicians are more likely to use smartphones in their studies and job, rendering them more exposed to these effects [5]. Mobile phone use has been associated with headaches, physiological irregularities, drowsiness, and tiredness [6]. Smartphones have given people control over how, when, and where they interact with others. Many smartphone users check their phones first thing in the morning and again before bed. Also, users have become accustomed to checking their phones hundreds of times every day. which can lead to “phone addiction” or obsessive phone use [7]. This study aimed to look at the trends in smartphone usage among medical students and see how they affect their academic performance.

According to recent research, technology plays an essential role in many facets of modern life and exposes students to diverse global knowledge [9]. Smartphones have become the choice for college students to communicate messages, complete assignments, and do research [10]. On the other hand, excessive smartphone usage or smartphone addiction may influence academic performance since students tend to use their cell phones for enjoyment rather than academic goals. The Choudhury and Tripathi study [11,12], which looked at the relationship between smartphone addiction and academic performance, found that high smartphone addiction reduces academic performance. The research also discovered an intrinsic association between internet access and smartphone addiction. Teens who have access to the Internet on their cell phones are more likely to communicate with their phones, which can lead to poor academic performance. Smartphone addicts may have poor time management skills since they spend most of their time on their phones and overlook other crucial aspects of their lives. Adolescents’ academic performance may be linked to poor time management caused by smartphone addiction. This is because phone addiction has been linked to poor performance [13]. A 2017 study reported that students nowadays rely on mobile phones for communication and social networking. It has also resulted in the acquisition of a psychological reliance on mobile phones, resulting in sleep deprivation and increased stress, which harms their academic performance. This study aims to see how mobile phone use affects sleep problems.

Furthermore, the findings revealed that the study’s 203 participants utilized smartphones and social media. The time spent on a mobile phone daily ranged from 5 minutes to 10 hours. Almost 72% of the participant’s population Intervention comparator outcome reported poor sleep quality, 66.5% had moderate weariness, and 14.8% experienced severe stress, with the majority (61%) using during the night hours. With smartphones, there was an essential link between poor sleep quality and academic performance [14]. Al-Osaimi et al. (2016., Al-Osaimi et al.) conducted a study to determine the prevalence of smartphone addiction among 2367 university students and the factors that lead to this addiction. The results revealed that 2.27% of the study sample spent more than 8 hours per day on their smartphones, while 75% spent less than 4 hours per day, and as a result of this use, the apps have at least one. Fell Per day, 43% of them sleep, feel energized, and 30% have unhealthy habits such as eating junk food, being overweight, and not exercising. The results showed a positive and substantial relationship between the degree of problematic use and the measure of academic success [15].

## Methodology

### Study design and settings

An observational cross-sectional study was conducted from July to September 2022 including students of the College of Medicine at King Saud University, Kingdom of Saudi Arabia. The data collection tool was structured and utilized an electronic survey.

### Ethics approval

The approval was taken from the College of Medicine, King Saud University 2022

### Data Collection Method/Data Source

A questionnaire was utilized to conduct this study. The questionnaire consists of two parts.1) section a consists of the general demographic characteristics of the respondents including current status, age, education, and gender. While 2) section b contains 4 subsections, consisting of 20 questions utilizing Google Forms. These sections are academic performance, the efficiency of the reaction, self-efficacy of a smartphone, and smartphone addiction.

The questionnaire was adopted from a previously published study developed, validated, and used by Mukhdoomi et al, Arooba and Farooqi, and others. Contacted to use it (2020) [8]. The survey link was distributed online.

The inclusion criteria for the study was the possession of a mobile phone among King Saud University College of Medicine willing to participate in the study.

A smartphone is a handheld electronic device that provides a connection to a cellular network and the internet.

### Power calculation

The sample size was calculated using a prevalence rate of 73.4% [18]. We used the prevalence at a 95% confidence interval, and the error rate is 5%. So, the sample size for this study would be 300 after adding a 10% attrition rate.

Ethics are the rules and standards for our research paper. Responses were kept confidential. Before participating in the study each student will sign a consent.

### Statistical Analysis

Descriptive statistics were calculated using means and standard deviations (SD). Student’s t-test and ANOVA. The Chi-square test was used to assess the dichotomous variables. All statistical analyses were conducted using the Statistical Package for the Social Sciences (SPSS) version 26 (Chicago, Illinois, USA).

## RESULTS

A total of 330 participants responded to the study questionnaire. The most common age range of study participants was 18-28 years, with 64.2%. Male participants represented 63%. The study population was Saudi citizens. Students from levels 3-4 accounted for 37%, followed by levels 5-6 at 30%. About 54.5% of the study’s income is less than 5000 riyals per month, and the unmarried participants were 55.8% (Table 1).

**Table 1:** Demographic characteristics of the study population

| Variables | Classification | Frequency | Percentage |
| --- | --- | --- | --- |
| Age | 18-28 | 212 | 64.2% |
|  | 29-39 | 114 | 34.5% |
|  | ≥ 40 | 4 | 1.2% |
| Gender | Male | 208 | 63.0% |
|  | Female | 122 | 37.0% |
| National | Saudi | 330 | 100.0% |
|  | Non- Saudi | 0 | 0.0% |
| Study stage | 1-2 | 82 | 24.8% |
|  | 3-4 | 122 | 37.0% |
|  | 5-6 | 99 | 30.0% |
|  | 7-8 | 27 | 8.2% |
| Income (SR. month) | ≤5000 | 180 | 54.5% |
|  | 5000 - 9000 | 128 | 38.8% |
|  | < 10,000 | 22 | 6.7% |
| Marital status | Married | 146 | 44.2% |
|  | Unmarried | 184 | 55.8% |
| Total |  | 330 | 100.0% |

It was found that using smartphones helped most students study more efficiently, 65%, while 35% were neutral and opposed. Regarding the use of smartphones on academic performance, about 60% of the participants agreed that their study performance had improved. Production of the training course and the enhancement of the effectiveness of their studies using smartphones, about 60% answered in agreement, while 40% were neutral. About 67% found that smartphones are useful in their studies, and the rest, 33%, disagree (Table 2).

**Table 2:** Scores of Smartphones and their impact on Academic Performance

| Questions Academic performance | Strongly Disagree (1) | Disagree (2) | Neutral (3) | Agree (4) | Strongly Agree (5) | Total |
| --- | --- | --- | --- | --- | --- | --- |
| Using a smartphone helps me to study more efficiently | 5 (1.5%) | 12 (3.6%) | 97 (29.4%) | 142 (43%) | 74 (22.4%) | 330 (100%) |
| Using a smartphone improves my performance in studying. | 5 (1.5%) | 9 (2.7%) | 113 (34.2%) | 136 (41.2%) | 67 (20.3%) | 330 (100%) |
| Using a smartphone increases my course work productivity. | 5 (1.5%) | 11 (3.3%) | 114 (34.5%) | 131 (39.7%) | 69 (20.9%) | 330 (100%) |
| Using a smartphone enhances my study effectiveness. | 6 (1.8%) | 7 (2.1%) | 111 (33.6%) | 143 (43.3%) | 63 (19.1%) | 330 (100%) |
| Overall, I find a smartphone useful in my studies. | 1 (0.3%) | 8 (2.4%) | 100 (30.3%) | 137 (41.5%) | 84 (25.5%) | 330 (100%) |

Regarding the efficiency of smartphone interaction in social relations with others, 70% agreed, and 30 were neutral and opposed. Speed of feedback was obtained using smartphones, 77% answered in agreement, and 22% were neutral. 44% agreed that they could interact with others using multiple tools. When dealing with others, regardless of their presence, 78% agreed, and the remaining 32% between neutrals and opponents. 70% of the participants reported y they can hold long conversations, and 25% are neutral (Table 3).

**Table 3:** Scores of Smartphones and their impact on Interaction Competency

| Questions Interaction Competency | Strongly Disagree (1) | Disagree (2) | Neutral (3) | Agree (4) | Strongly Agree (5) | Total |
| --- | --- | --- | --- | --- | --- | --- |
| With a smartphone, I can maintain social relationships with others. | 5 (1.5%) | 9 (2.7%) | 77 (23.3%) | 135 (40.9%) | 104 (31.5%) | 330 (100%) |
| With a smartphone, I can get feedback quickly. | 0 (0%) | 3 (0.9%) | 73 (22.1%) | 142 (43%) | 112 (33.9%) | 330 (100%) |
| With a smartphone, I can interact with others using multiple tools. | 2 (0.6%) | 7 (2.1%) | 81 (24.5%) | 147 (44.5%) | 93 (28.2%) | 330 (100%) |
| With a smartphone, I can interact with others no matter where they are. | 2 (0.6%) | 3 (0.9%) | 68 (20.6%) | 131 (39.7%) | 126 (38.2%) | 330 (100%) |
| With a smartphone, I can easily have a longer conversation with others. | 9 (2.7%) | 4 (1.2%) | 85 (25.8%) | 132 (40%) | 100 (30.3%) | 330 (100%) |

Our study found that 69.1% use smartphones to take exams and register for courses, and 31% are neutral. The use of smartphones for training and education sites was in favor with 69.4%, while 30% were neutral. Almost 69% agreed they could complete tasks and presentations, while the remaining 31% were neutral and disagreed. When searching for information using smartphones, we found 87.8% agree, and 11% are neutral (Table 4).

**Table 4:**
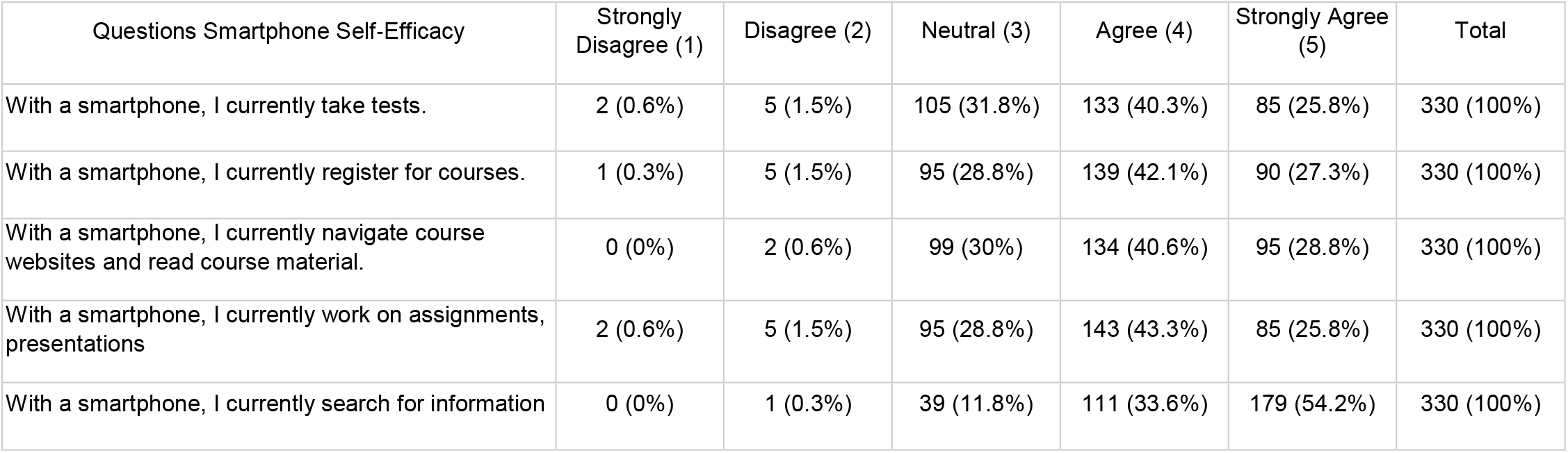
Scores of Smartphones and their impact on Self-Efficacy

Our study indicates that 81.2% spend more than 5 hours on their smartphone. About 71.2% of the respondents answered that it is difficult for them to turn off their smartphones and go about their day without a smartphone. 58.8% unconsciously use their smartphone to check calls and messages. Phone bill spending was at most 23%, as 76% spent less on their phones than on clothes or food (Figure 5).

**Figure 5:**
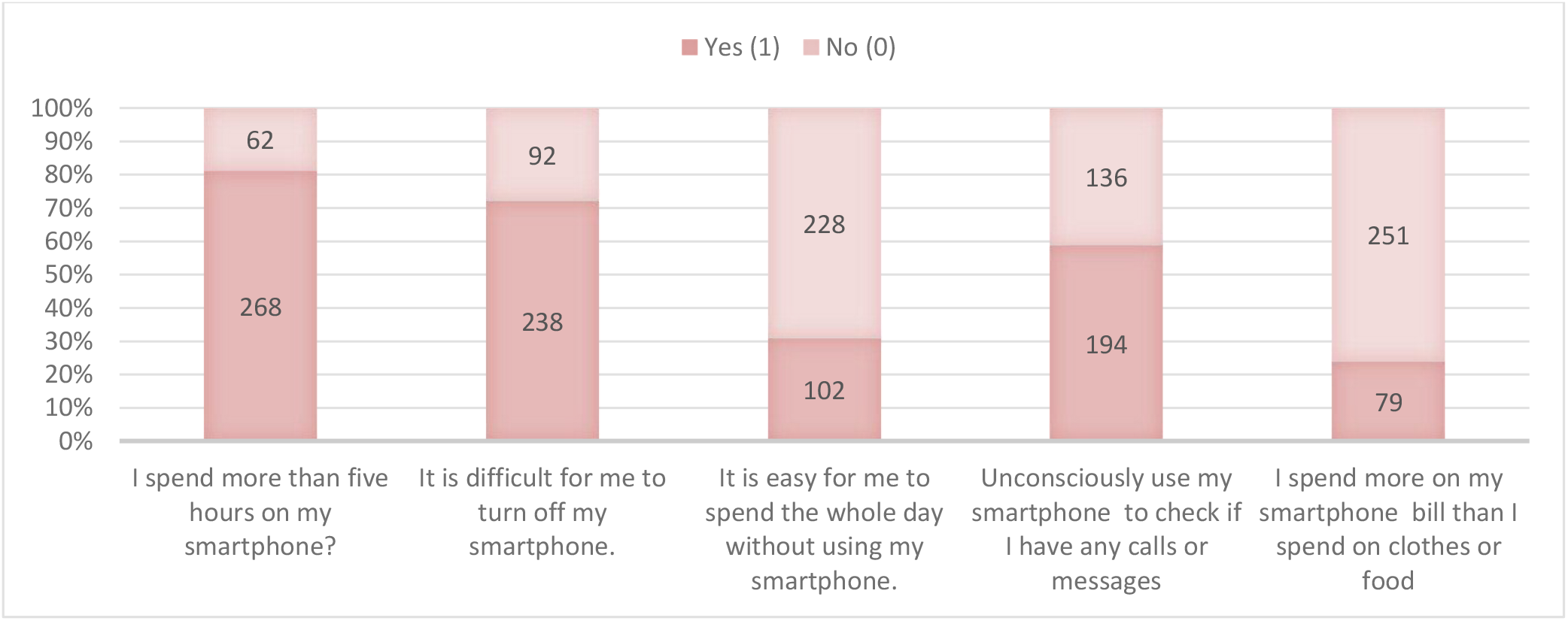
Scores of Smartphones addiction

## Discussion

Nearly all Saudi college students use smartphones, especially those studying medicine. Extreme smartphone usage has largely disappeared and is now categorized as addictive behavior[17]. In our study, we assessed the use of smartphones and their relationship to academic performance at the College of Medicine at King Saud University, Kingdom of Saudi Arabia, in 2022 was the main emphasis of this study. This description of the results makes links with the study’s goals and may be supported by previous literature reviews.

Our statistics and the study of Aznar-Daz et al. are not relatively consistent. The study findings show that more male medical students than female students utilized smartphones. According to Chen et al. (2017), more females than males are present, which contradicts our findings. As for age groups, those between the ages of 18 and 28 (64.2%) and 29 and 39 (34.5%) students had the highest phone addiction rates. However, Aznar-Daz et al. (2019) acknowledged that phone addicts were mainly above 30 and in the age range of 18 to 21. In addition, our research supported this study’s finding that one of the factors contributing to smartphone addiction is work.

Understanding how smartphones are used for academic achievement was the initial research objective. According to the research, 2.7% of students disagree with the 41.2% who claim that smartphones help their academic achievement. The widespread use of smartphones demonstrates the benefits the technology offers its consumers. On the other hand, a 2017 study revealed that developing a psychological reliance on mobile devices causes sleep loss and elevated stress, which has a detrimental effect on students’ academic performance. After looking at the data in Table 2, it was shown that the majority of students believed smartphones to be very beneficial to them since they enhance their performance (41.2%), increase their productivity (39.7%), and are generally helpful for them in their studies (41.5%). Some students did disagree, although their numbers were relatively few. On the contrary, Mohammadi et al. (2020) discovered that despite their advantages, phones were not accepted by students.

The second research objective was to depict interaction competency, which is displayed in table 3, with 40.9% of participants agreeing and 2.7% disagreeing that smartphones are helpful for social interaction. However, a 2017 study found that individuals used their phones far more than average and, as a result, suffered from fatigue, extreme stress, and sleep disturbances. The majority of participants consider smartphones to be socially compelling because users can have lengthy conversations (40%), immediately provide feedback (43%), maintain relationships on social media (40.9%), and communicate in a variety of ways (44.5%). Ihm (2018) further acknowledged that although utilizing phones promoted social support among students, addiction could develop if phones were used excessively.

Explaining the self-efficacy of smartphones, as demonstrated in table 4, was the third research objective. 42.1% of respondents utilized smartphones for course-taking, although only 1.5% did not. Choudhury and Tripathi (2018) established a direct link between smartphone addiction and internet access, resulting in subpar academic performance. Students overwhelmingly stated that smartphones made it easier for them to take tests (40.3%), read course material (40.6%), make presentations (43.3%), and search and browse online (33.6%). The findings of our study are consistent with Mohammadi et al. (2020), who noted that smartphones draw students’ attention using software applications like Word, Excel, and PowerPoint.

As demonstrated in table 5, the fourth research objective was to assess smartphone addiction. Results indicate that 41.2% of respondents did not check their smartphones for calls or messages, while 58.8% did so unintentionally. The findings indicated that the students had developed a dependence on their smartphones since they admitted to using them frequently (81.2%), being unable to put them away (72.1%), and finding it difficult to live without them (69.1%). According to Choudhury and Tripathi (2018), adolescent students’ academic success may be affected by poor time management brought on by smartphone addiction.

Study limitations and recommendations for future Studies

The use of smartphones among medical students in Saudi Arabia has grown in recent years due to the public’s interest in the technology. Smartphones are becoming a daily need for university students, which is another factor contributing to the high usage level. However, there are several gaps and limitations in this study. Although this study was restricted to a particular campus, the implications of smartphone use by university students extend to all higher education institutions. It is appropriate for a study of this kind to be conducted at other academic institutions.

Further research should be conducted to investigate the influence of smartphones on university students’ continuous concentration in their academics and college attendance. There has to be research on how students write academically and smartphone usage in developing nations. A study on how smartphones affect students’ cultural values and worldviews should be conducted, among other things. Due to a lack of time and resources, this study could not cover these topics.

## Conclusion

Our study found a significant correlation between phone addiction and a drop in academic performance. Despite its attractiveness, smartphone addiction is a time waster for students that might disrupt their sleep and causes stress. The wise use of smartphones in e-learning services is promising, so it is vital to create a comprehensive plan that guides students toward productive use

## Data Availability

The inclusion criteria for the study was the possession of a mobile phone among King
Saud University College of Medicine willing to participate.

https://docs.google.com/forms/d/1__1Kss3b7VGCmfVHFvXOqMRCuZxOzfWdJYo7e-c6zbU/prefill

